# Moving Singing for Lung Health online: experience from a randomised controlled trial

**DOI:** 10.1101/2020.08.07.20170050

**Authors:** Keir EJ Philip, Adam Lewis, Edmund Jeffery, Sara Buttery, Phoene Cave, Daniele Cristiano, Adam Lound, Karen Taylor, William D-C Man, Daisy Fancourt, Michael I Polkey, Nicholas S Hopkinson

**Affiliations:** National Heart and Lung Institute, Imperial College London, London, United Kingdom; Department of Health Sciences, Brunel University, London, United Kingdom; Royal Brompton and Harefield NHS Foundation Trust, London, United Kingdom; Imperial College Healthcare NHS Foundation Trust, London, United Kingdom; Department of Behavioural Science and Health, University College London, London, United Kingdom

**Keywords:** COPD, Singing, arts in health

## Abstract

**Introduction:** Singing for Lung Health (SLH) is a popular arts-in-health activity for people with long-term respiratory conditions, which participants report provides biopsychosocial benefits, however research on impact is limited. The ‘SHIELD trial’, a randomised controlled, single (assessor) blind, trial of 12 weeks SLH vs usual care for people with Chronic Obstructive Pulmonary Disease (COPD) (n=120) was set-up to help to address this. The first group started face-to-face (5 sessions) before changing to online delivery (7 sessions) due to COVID-19 related physical distancing measures. As such, the experience of this group is here reported as a pilot study to inform further research in this area.

**Methods:** We conducted semi-structured interviews and thematic analysis regarding barriers, facilitators and key considerations regarding transitioning from face-to-face to online delivery. Pilot quantitative outcomes include attendance, pre and post measures of quality of life and disease impact (SF-36, CAT score), breathlessness (MRC breathlessness scale, Dyspnoea-12), depression (PHQ9), anxiety (GAD-7), balance confidence (ABC scale) and physical activity (clinical visit PROactive physical activity in COPD tool, combining subjective rating and actigraphy).

**Results:** Attendance was 69% overall, (90% of the face-to-face sessions, 53% online sessions). Analysis of semi-structured interviews identified three themes regarding participation in SLH delivered face-to-face and online, these where 1) perceived benefits; 2) digital barriers (online); 3) digital facilitators (online). Findings were summarised into key considerations for optimising transitioning singing groups from face-to-face to online delivery. Pilot quantitative data suggested possible improvements in depression (treatment effect −4.78, p= 0.0487, MCID 5) balance confidence (treatment effect +17.21, p=0.0383, MCID 14.2), and anxiety (treatment effect −2.22, p=0.0659, MCID 2).

**Discussion:** This study identifies key considerations regarding the adaptation of SLH from face-to-face to online delivery. Pilot data suggest online group singing for people with COPD may deliver benefits related to reducing depression and anxiety, and improved balance confidence.

**KEY MESSAGES:** *What is the key question?:* Can Singing for Lung Health (SLH) be delivered online for people with COPD? And if so, what are the practical issues and how does the experience compare with face-to-face participation?

*What is the bottom line?:* SLH appears safe and enjoyable both face-to-face and online. Access barriers for online sessions included digital access and literacy. However increasing access to those previous unable to physically access sessions is also important. In this pilot, depression, anxiety and balance confidence appear to show improvements related to participation in a SLH group that transitioned from face-to-face to online delivery.

*Why read on?:* To our knowledge this is the first study to assess health impacts of online group singing sessions. Given the physical distancing measures required by the response to COVID-19, there is a need for singing groups and other similar interventions to be delivered online such as pulmonary rehabilitation. This study helps to inform this and future research in the area.

## Introduction

Many people with Chronic Respiratory Disease (CRD) remain highly symptomatic despite optimal pharmacological treatment. Symptoms including exercise limitation, shortness of breath, and depression are common^1-3^. These can be compounded by social isolation and loneliness, which have been shown to be important to respiratory health outcomes^4^. Group singing is a common practice in most societies globally and has been shown to have health and wellbeing benefits for people with living with long-term health conditions and the wider general public^5-7^. There is increasing interest in arts-in-health interventions for people with chronic health conditions from patients through to government level^8^. Singing for Lung Health (SLH) is a popular group singing programme specifically developed for people with CRD. Small scale trials and qualitative studies suggest SLH has a range of biopsychosocial benefits for participants^9 10^, however there is a lack of larger, longer term, randomised control trials (RCT) regarding the impacts of this intervention^11^. The *‘Singing for Lung Health: Improving Experiences of Lung Disease (SHIELD) trial’* was set up to help to address this gap, planning to randomise 120 individuals with COPD to participation in 12 weeks of group singing or usual care.

During the current COVID-19 pandemic, physical distancing measures aimed at reducing SARS-CoV-2 transmission have led to profound social adaptations and disruption. Group activities, particularly involving people with long term health problems who are especially vulnerable to COVID-19, have in most cases been suspended including pulmonary rehabilitation programmes - one of the highest value interventions for people with respiratory disease^12 13^. Similarly, there are particular concerns that group singing could be a high risk activity regarding viral transmission, however research is currently limited^14^.

This context has driven interest in the implementation, and ongoing development, of online approaches that attempt to reproduce the social, psychological and physical effects of singing, dance and more established interventions such as pulmonary rehabiliation^15 16^. Such approaches are especially important for people with lung conditions as even prior to the COVID-19 pandemic access to these interventions was inadequate^17 18^. Furthermore, measures to reduce risk of COVID-19 in this group appear to be causing substantial disruption to care and access to health-services, with high levels of anxiety and loneliness being reported^19 20^.

The first group of participants in the SHIELD trial, which began in February 2020, initially met face-to-face but the delivery of the intervention changed to an online format which is likely to remain necessary for the foreseeable future. We decided that this transitional group should be reported as a pilot study. This was firstly because research on the health and wellbeing impacts of online singing groups is lacking so the results could guide both the further delivery of the SHIELD trial and the design of other studies. Secondly, this would provide useful information from individuals who had experience of both face-to-face and online activities and could therefore enable a comparison of the two types of intervention experience.

## Methods

### Trial design and oversight

The SHIELD Trial was prospectively registered at clinicaltrial.gov (NCT04034212). The current analysis was defined as an amendment to the initial trial registration when the delivery of singing moved from face-to-face to online. Ethical approval for the SHIELD trial was granted by the National Health Service Health Research Authority, Stanmore REC (19/LO/0418).

### Participants

The first group of 18 participants in the SHIELD trial were recruited from a specialist COPD clinic at the Royal Brompton Hospital London and lists of previous research participants who had given consent to be contacted regarding research (Figure 1). Chronic Obstructive Pulmonary Disease (COPD) diagnosis was confirmed by spirometry. Exclusion Criteria included pulmonary rehabilitation less than 4 months ago (potential to impact outcome measures due to potential overlap of certain impacts), inability to take part in singing sessions due to comorbidity (e.g. life limiting illness, cognitive impairment), and previous participation in SLH classes. Given the requirement for the original protocol of weekly in-person attendance, from the list of potential participants, people living within a 1-hour journey of the hospital (estimated using google maps) were contacted first. All participants provided written informed consent after reading the Participant Information Sheet (PIS) and being given the opportunity to ask questions. Transport costs related to the assessment visit were reimbursed, but no payments were made for participation.

**Figure 1:**
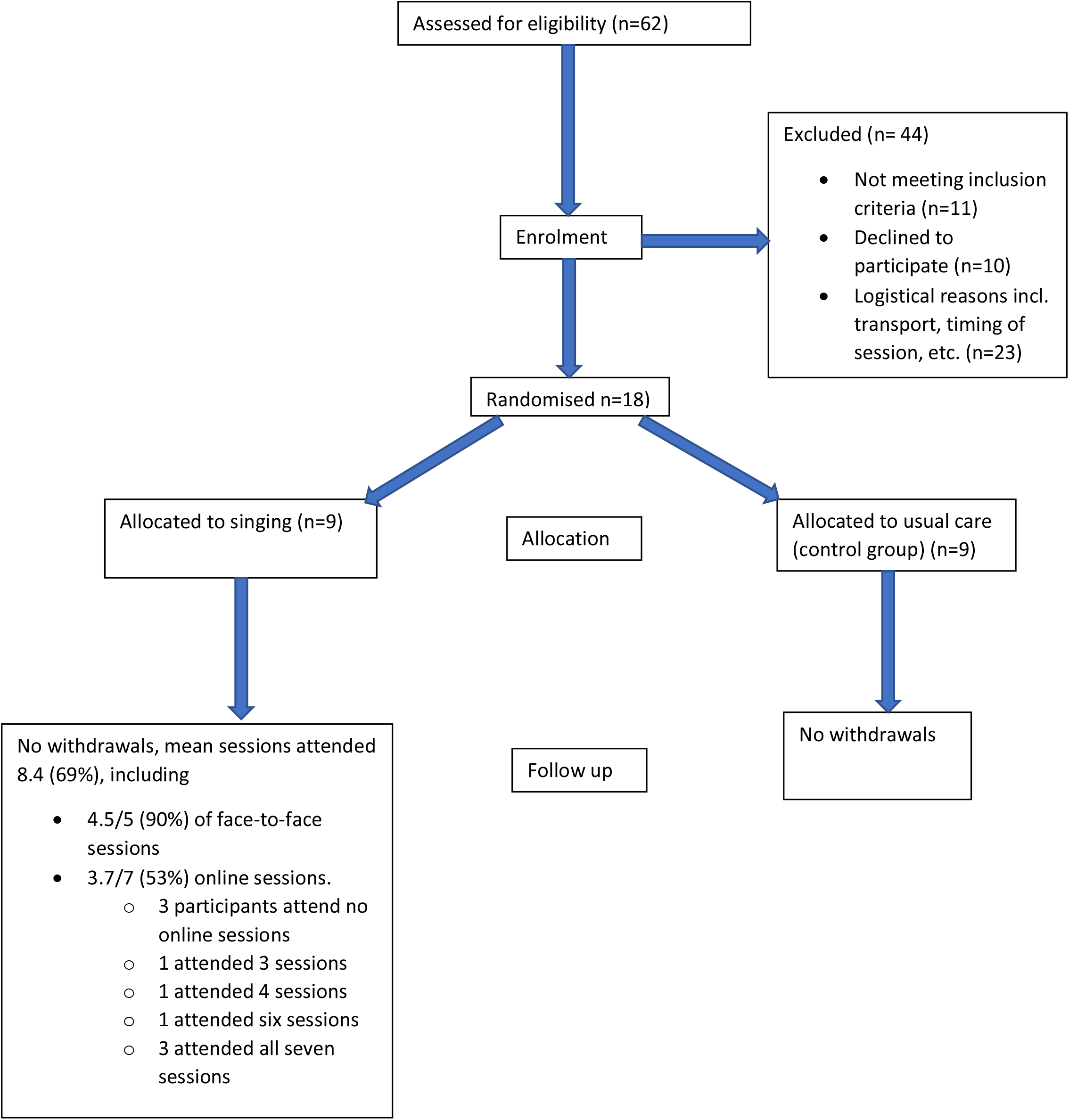
Moving Singing for Lung Health online: experience from a randomised controlled trial: Consort diagram.

### Patient and Public Involvement

The Royal Brompton Clinical Research Facilities patient expert and lay person research panel, and the Royal Brompton ‘Breathe Easy’ Group, reviewed the study proposal, provided thoughts and suggested improvements which improved the study design and materials prior to Research Ethics Committee review. In particular, the choice of a quality of life primary outcome measure was well received. The PIS was clarified including reducing use of specialist terms and specifying which study related costs would be reimbursed. Overall, the study was very well received by these patient and public representatives who saw clear value in the patient centred focus of the study.

### Interventions

The intervention arm (SLH) received 12, once weekly, hour long, SLH sessions. The study control arm received usual care (UC) (no specific additional intervention above those which the person usually engages with). The specific content and structure of the singing sessions has been described elsewhere^10^, but briefly, each session consisted of a physical warm up, breathing exercises, vocal warms ups, songs, and a cool down. The sessions were led by Edmund Jeffery, a professional singing teacher with four years’ experience leading SLH groups. The SLH participants also received a CD of singing exercises and were encouraged to practice between sessions.

The first five, weekly sessions were delivered face-to-face as originally planned at the Royal Brompton Hospital, London, UK. And started on the 10/02/2020. These were halted due to the developing COVID-19 situation in the UK for 2 weeks to develop the online delivery of sessions. This process included discussion within the SLH provider network regarding suitable content, potential barriers and facilitators, and trial sessions with experienced leaders to help address unidentified challenges. The online SLH format which was developed was then used to deliver the final 7 sessions via a video conferencing application (Zoom Video Communications Inc. “Zoom”).

### Baseline assessment, randomisation and blinding

Assessments took place at the Royal Brompton Hospital (London, UK). Following a screening visit, written informed consent was taken, followed by a structured clinical history, and confirmation of COPD diagnosis by spirometry, and baseline assessment of outcome measures. Participants were randomised (1:1) using computer-generated block randomisation lists (Sealed Envelope™) block size 4, stratified by Medical Research Council (MRC) breathlessness grade and previous participation in pulmonary rehabilitation. The outcome assessors were blind to intervention allocation. Blinding of participants was not possible, and they were informed of their allocation by the unblinded researcher responsible for randomisation (SB). Unblinding of assessors took place only after all outcome measure data had been recorded.

### Outcome measures and assessment

Primary outcome measures were change in the Short Form 36 Physical and Mental Component Scores (MCS and PCS respectively) using the oblique scoring method^21^, with sub-scales provided to aid interpretation. Secondary outcomes included change in SF-36 subscales, balance confidence (ABC scale), anxiety (GAD-7), depression (PHQ-9), COPD assessment test (CAT) score, breathlessness (MRC dyspnoea score and Dyspnoea-12). Patient experience of physical activity was assessed using the clinical visit PROactive physical activity in COPD tool (cPPAC). The cPPAC combines questionnaire and seven days of actigraphy measures to produce two domains, amount and difficulty^22 23^.

The original SHIELD protocol included physical capacity and performance testing using the six-minute walk test (6MWT) and the short physical performance battery (SPPB). However, these were only conducted at baseline as repeating these measures was not possible due to COVID-19 related physical distancing measures.

Outcomes measures were repeated after 14 weeks (12 weeks intervention plus 2-week pause for adaptation of delivery after week 5 of intervention). Baseline assessments were completed within 4 weeks prior to the intervention arms first singing session. Follow-up assessments were completed within 4 weeks of study completion by participants at home, with activity monitors and questionnaires returned by post. Any missing data (unanswered questions) were addressed by telephoning the participant.

Semi-structured qualitative feedback interviews were conducted on the phone (by KP who has training and experience in qualitative research techniques) with the SLH participants following unblinding of researchers, focusing on overall experience of intervention, positives, negatives, and barriers and facilitators to participation (as covered with an approved amendment to the Research Ethics Committee application). Notes were made during the call and a template response form was completed immediately after each call to record participant responses and interviewer reflections. Deductive thematic analysis was used^24^. Comparison of experiences of face-to-face and online groups was also sought.

## Statistical analysis

The power calculation for the SHIELD trial required 120 participants to show a clinically important difference in the primary outcome (SF-36). For this revised pilot study we present p values for independent sample t-tests between intervention and control groups. Analyses were carried out using Stata 14 (StataCorp, TX) on an intention to treat basis.

## Results

The intervention and control groups were well-matched at baseline (Table 1). Only GAD-7 anxiety score was of borderline statistical significance for between group difference at baseline (p 0.057), mean values of 6.33 (UC), 1.88 (SLH). No serious adverse events were reported. No participants withdrew from the study. However, difficulties with attendance of the online sessions is described below.

**Table 1:** Baseline characteristics:

|  | Singing for Lung Health (SLH) (n=9) | Usual Care (UC) (n=9) | p-value |
| --- | --- | --- | --- |
| Age | 71 (69,78) | 70 (69,72) | 0.6 |
| Female | 3 (33%) | 6 (66%) | 0.2 |
| BMI | 19 (19.6, 23.4) | 25 (23.3, 30) | 0.5 |
| FEV<sub>1</sub> % predicted | 40 (37, 77) | 33 (30, 48) | 0.1 |
| Oxygen therapy | 1 (11%) | 2 (22%) | 0.6 |
| Pack years smoked | 38 (23, 55) | 40 (15, 53) | 0.7 |
| Falls in last year | 1 (11%) | 1 (11%) | 1.0 |
| Baseline 6MWD | 405 (340, 537) | 395 (309, 419) | 0.3 |
| SPPB total | 10 (8, 11) | 10 (8, 10) | 0.7 |
| Pre-study expectation of SLH health impact<br>(0 'no impact' to 10 'large impact') | 4 (3, 4) | 2 (2,5) | 1.0 |
| SF-36 Physical Component Score | 40.0 (32.7, 47.0) | 34.5 (33.3, 36.9) | 0.3 |
| SF-36 Mental Component Score | 44.6 (41.8, 49.2) | 44.0 (34.3, 46.9) | 0.2 |
| SF-36 Physical function | 40 (25, 65) | 35 (20, 50) | 0.6 |
| SF-36 Role limitation, physical | 25 (0, 50) | 0 (0, 50) | 0.2 |
| SF-36 Role limitation, emotional | 66.7 (33.3, 66.7) | 33.3 (0,66.7) | 0.1 |
| SF-36 energy | 50 (25, 55) | 50 (40, 50) | 0.8 |
| SF-36 emotional well being | 80 (68, 80) | 64 (60, 68) | 0.2 |
| SF-36 social functioning | 87.5 (62.5, 100) | 62.5 (50, 100) | 0.2 |
| SF-36 pain | 67.5 (45, 87.5) | 77.7 (42.5, 77.5) | 0.7 |
| SF36 general health | 30 (5, 30) | 20 (10, 30) | 0.9 |
| SF-36 health change | 25 (25, 50) | 50 (25, 50) | 0.6 |
| Depression (PHQ-9) | 5 (3, 8) | 6 (5,14) | 0.2 |
| Anxiety (GAD-7) | 2 (1, 2) | 5 (0, 11) | 0.1 |
| ABC scale score | 78.8 (50.6, 90.6) | 76.3 (64.4, 91.3) | 0.7 |
| Dyspnoea 12 | 11 (3, 15) | 13 (8, 20) | 0.3 |
| MRC dyspnoea score | 3 (2, 4) | 3 (2, 4) | 0.9 |
| CAT score | 17 (13, 22) | 21 (17, 27) | 0.5 |
| PROactive difficulty* | 72 (60, 75) | 68 (56, 75) | 0.6 |
| PROactive amount** | 57 (44, 65) | 48 (44, 52) | 0.3 |
| PROactive total*** | 61 (59, 72) | 58 (53, 60) | 0.2 |
| Daily step count | 2717 (1870, 4871) | 2566 (1213, 3119) | 0.4 |

\*scale of 0 (high level difficulty) to 100 (low level of difficulty); \*\* scale of 0 (low amount) to 100 high amount; \*\*\* total score calculated as mean of PROactive difficulty and PROactive amount (0-100)
Data shown are median (IQR) or number (%). P values are for t tests test between groups. BMI, body mass index; 6MWD, six minute walk test distance; SPPB, short physical performance battery; Pre-study perception of SLH impact, participants asked to rate on scale of 0 to 10 expected impact of singing on improving health with 0 is no impact, and 10 very large improvement in health; SF-16,
*Short Form 36 Health Survey; PHQ-9, Patient Health Questionnaire-9; GAD-7, Generalised Anxiety Disorder-7 questionnaire; ABC score, Activity specific Balance Confidence scale; MRC, Medical Research Council; CAT, COPD Assessment Test; PROactive, Patient Reported Outcome measure for physical activity in COPD.*

### Attendance

For the SLH arm, mean number of sessions attended was 8.4 (69%) of the 12 total sessions. This included a mean of 4.5 (90%) of the 5 face-to-face sessions, and 3.7 (53%) of 7 online sessions. Three of the participants did not attend the online sessions at all, three attended all seven sessions, with the remaining three participants attending three, four and six sessions.

### Effect on intervention

Change from baseline was compared between the control and SLH groups in Table 2. These are presented for information, but cannot, of course, be used to make any confident inference about the effectiveness or otherwise of the intervention given the limited sample size compared to that for which the study was powered. Comparing singers with non-singers there were statistically significant improvements in the PHQ-9 depression score (treatment effect −4.78, p= 0.049, MCID 5) and ABC scale for balance confidence (treatment effect +17.21, p=0.038, MCID 14.2). There was also a suggestion of benefit regarding anxiety levels (GAD-7) in the singers of borderline statistical significance (treatment effect 2.22, p=0.066, MCID 2).

**Table 2:** Comparison of change in outcome measures between study arms.

|  | Singing for Lung Health (SLH) (n=9) | Usual Care (UC) (n=9) | Treatment effect | p-value |
| --- | --- | --- | --- | --- |
| SF-36 Physical Component Score (PCS) (Oblique calc) | -1.66 | -0.389 | -1.27 | 0.67 |
| SF-36 Mental Component Score (MCS) | -0.367 | -4.30 | +3.93 | 0.15 |
| SF-36 Physical function | -1.11 | 2.77 | -3.89 | 0.79 |
| SF-36 Role limitation, physical | -16.67 | -2.78 | -13.89 | 0.84 |
| SF-36 Role limitation, emotional | -7.41 | -33.33 | +25.92 | 0.10 |
| SF-36 energy | 6.11 | -1.11 | +7.22 | 0.19 |
| SF-36 emotional wellbeing | 2.22 | -6.22 | +8.44 | 0.14 |
| SF-36 social functioning | -6.95 | -15.28 | +8.33 | 0.26 |
| SF-36 pain | 0.28 | 5.78 | -5.50 | 0.68 |
| SF36 general health | 0 | 1.67 | -1.67 | 0.62 |
| SF-36 health change over last year | 5.56 | -13.89 | +19.44 | **0.026** |
| Depression (PHQ-9) (MCID 5<sup>25</sup>) | -1.44 | +3.33 | -4.78 | **0.049** |
| Anxiety (GAD-7) (MCID 2<sup>26</sup>) | 0.111 | 2.33 | -2.22 | 0.066 |
| ABC scale score (MCID 14.2<sup>27</sup>) | 6.03 | -11.18 | +17.21 | **0.038** |
| Dyspnoea 12 | -0.445 | 0.445 | -0.889 | 0.39 |
| MRC dyspnoea score | 0.222 | 0.111 | +0.111 | 0.63 |
| CAT score | -1.44 | 2.22 | -3.67 | 0.098 |
| PROactive Difficulty* | 0.889 | -1.12 | +2.00 | 0.362 |
| PROactive amount** | -20.22 | -6.89 | -13.33 | 0.95 |
| PROactive total*** | -9.67 | -4.00 | -5.67 | 0.92 |
| Daily step count | +129 | -1408 | +1537 | 0.13 |

*P values are independent sample t-tests. \*change in the scale of 0 (high level difficulty) to 100 (low level of difficulty); \*\*change in the scale of 0 (low amount) to 100 high amount; \*\*\*change in the scale of the total score calculated as mean of PROactive difficulty and PROactive amount (0-100).*

**Table 3:** Considerations for online vs face-to-face delivery of Singing for Lung Health.

|  | Face-to-face | Online |
| --- | --- | --- |
| **Access: Physical** | More challenging<br>Geographically local SLH sessions<br>Transport requirement<br>Financial and time costs<br>Availability and accessibility in context of lung condition and symptoms | Private physical space still required for participant |
| **Access: Online** | Limited or no requirement<br>Can be used for session organisation | Computer/device and internet access required.<br>Overcomes multiple face-to-face physical barriers |
| **Digital Literacy** | Not required | Required<br>With support can be minimal<br>Could help build skills/confidence facilitating access to other online services |
| **Infection/health risks** | No risk of cross infection from participants or infection during transport to/from session | Not supervised in person<br>Contact details of next of kin advisable for potential issues |
| **Social experience** | Very effective and important to participants | Good (perhaps less than face-to-face)<br>Building rapport more challenging, especially if have never met in person (new groups/individuals)<br>Specific content to address advised |
| **Personal experience** | Presence of peers can be both supportive and slightly intimidating depending on individual and group dynamic | The required technological skills can induce mild anxiety if participant not confident/experienced. |
| **Physical engagement** | Important aspect of session<br>Journey to/from session also valuable physical activity for some | More challenging than face-to-face<br>Requires specific consideration and promotion |
| **Facilitator experience** | Easier to gauge participant emotions and group dynamic | More challenging to assess if appropriate techniques are being used by participants |

**Table 4:** Practical issues transferring face-to-face to online.

|  |  |
| --- | --- |
| **Informing participants** | People are generally understanding of current requirement for physically distanced sessions. |
| **Joining online** | Participants may need support with digital access.<br>Friends and family a good source of support.<br>Dedicated 'set up sessions' could help in which session leader talks the participant through setting up, 1 to 1, separately and in advance of the SLH session. |
| **Physical space** | Clean, tidy and free from trip hazards |
| **Sound** | Speakers advisable for better volume and sound quality |
| **Feedback** | Integration of formal feedback vital to facilitate responsive and participant appropriate sessions.<br>More immediate feedback required from session leader as participants are not able to hear each other sing. |
| **Maintaining/building relationships** | Introductions.<br>General catch up/social.<br>Rapport building activities. |
| **Session content** | Cannon and multi-harmonies (live) – difficult.<br>Pre-recorded content to facilitate multipart songs possible, but complicated.<br>Focusing on what works best in each method of delivery more important than trying to exactly emulate face-to-face sessions online.<br>Discussion between singing leaders using similar participant groups and swapping ‘best practice’ from experience works well. |
| **Keep up to date** | The evidence base in this area is evolving, so ongoing review of relevant research and guidelines is important, including related activities such as pulmonary rehabilitation<sup>28</sup> |

### Physical activity

The PROactive total score suggested that the SLH group did worse than the control group regarding change in the composite measure of amount and difficulty of physical activity. This was primarily driven by the larger subjective reductions in the amount of physical activity in the SLH group during the post-intervention follow-up assessment. However, this was discordant with objective assessment, the mean change in step count was +1537 higher in the SLH group, compared with the control group. The small sample, and “lock-down” related to COVID-19 physical distancing, limits confidence interpreting the significance of these findings.

### Participant experience

Deductive analysis identified three key themes regarding the SLH participants’ experience 1) perceived benefits; 2) digital barriers; and 3) digital facilitators.

#### Perceived benefits

All SLH participants reported greatly enjoying participation while session delivery was face-to-face. The online sessions were enjoyed by the majority of participants, though all stated an overall preference for face-to-face sessions. Participants reported it *‘helps your breathing’* and *‘Certainly my breathing is better now than good than before’* especially in relation to the breath control exercises and breathing techniques. Improvements to mood and enjoyment of the social aspects were frequently reported. There was also a perception that the singing had contributed to other types of physical activity *‘the singing has contributed to my exercise levels’*.

#### Digital barriers

Barriers relating to online delivery mostly related to technical difficulties. The majority experienced some form of technical difficulties; only those who regularly used online conferencing tools had no issues. Of those that did not attend at all online, one did not have a computer and one did not have a functioning internet connection. The third participant who did not participate online chose not to as they felt making noise would not be considerate of their neighbours, given the limited sound insulation, and ongoing ‘lockdown’ measures for all the residents of his building. Some participants were able to ask friends or family members to help with overcoming technological challenges, which worked well. However due to the “lock-down” and social shielding, this was not possible for all the participants.

Online delivery was also felt to be less personal, as interaction between participants was more challenging *‘meeting at the Brompton was much more engaging. It’s so nice to sing together as a group’*. One participant stated that *‘Physically demanding things are better done in a group’*, with group motivation more palpable in person than online. Even though face-to-face was preferred, online deliver was still seen as being extremely valuable *‘Even on online, it’s an up-lifting thing to do for mental health. We spent quite a lot time laughing. Singing as a group is special.’* Other aspects of the sessions were also noted to be lost when adapting the sessions to online. For example ‘*singing in canon (a compositional technique in music)’*, which was thoroughly enjoyed in person, were not technically possible during sessions delivered online.

It was felt that by having started the sessions face-to-face a degree of rapport had been built between participants, and with the singing leader, that helped with the transition online. It was suggested that it might be more difficult establishing relationships between group members if the groups commenced online with no prior meeting in person.

#### Digital facilitators

On certain points, online delivery was perceived as having benefits over in person sessions. One participant highlighted that ease of attendance enabled her to attend when she was not feeling 100% well and would not have attended in person had she needed to physically transport herself to the singing session. Another participant felt that by being online he was less self-conscious singing with the other participants as they would not be able to hear his voice. Multiple participants highlighted that online delivery overcame many of the barriers related to physically accessing face-to-face participation including geographical location of sessions, using transport with high symptoms burden, and currently infection risks.

## Discussion

To our knowledge, this is the first study to assess the practical delivery of an online group singing intervention for people with respiratory disease intended to improve health and wellbeing. This transition from face-to-face to online delivery was forced on us by the COVID-19 pandemic, but provides useful information about how this can be done and allowed us to gain insights from people who had experienced both formats of delivery.

Key findings include that SLH delivered online was viewed as enjoyable and holistically beneficial to health, though face-to-face was generally preferred. Technological difficulties prevented some people from participation in online sessions, which were also felt to be less personal as social interaction was more challenging. The pilot data suggest that group singing for people with COPD, adapted to online delivery, may still deliver benefits related to reducing depression and anxiety, and improved balance confidence.

These findings are broadly supportive of other related studies. However, these findings should be interpreted within the context of COVID-19 related physical distancing and ‘shielding’ measures. In a small (n=28) RCT of a six week course of twice weekly face-to-face SLH, Lord et al (2010)^29^ also found improvements related to anxiety, and qualitative research is has reported similar findings to the current study in relation to perceived impact on health and wellbeing^30^. Previous studies on SLH for people with COPD have suggested improvements in quality of life^29 31 32^, which was not seen here, though it is not clear if this related to the small sample size. Furthermore, our findings echo those of a study comparing the experiences of participants in a virtual choir, with those in a live choir, found the two types of experience provide very similar emotional benefits, though differences in how ‘present’ participants felt^33^.

The qualitative data identified specific barriers and facilitators related to the different formats of delivery, which helps to explain the attendance data. The consort diagram highlights that many (n=23) potential participants declined due to issues with physically accessing the face-to-face sessions, mainly related to viewing public transport as too challenging given their health condition. However, online delivery demonstrated good potential to overcome physical distance as a barrier to access. As highlighted in the interview feedback, during the online section of the study, there were days when individuals participated but felt they would not have felt well enough to come in person if the sessions were still being held at the hospital. Regarding the online sessions, difficulties with digital literacy and digital access presented barriers, in some cases preventing ongoing participation in those who stated they deeply enjoyed the face-to-face sessions.

The consort diagram may have been different if the methods of delivery had been known from the start. Some people who declined participation due to difficulties physically accessing the hospital may have participated. However, those for whom digital delivery poses barriers may have declined. Clearly digital access is a vital consideration to address to overcome this potential barrier to participation. Additionally, participant rapport building appears to be an area requiring particular consideration.

Some limitations to this study are important to discuss. Firstly, adaptation of the methods during the study was not ideal. That said, it has provided a unique opportunity to gain insights into the transition of singing groups to online delivery which have, by necessity, become widespread. Secondly, given the novelty of the online delivery, the session content and technical considerations are likely to develop overtime with experience, which could alter the relevance of the current findings to future sessions. However, this is not necessarily a weakness, as the current findings provide useful results on which to base these developments. Thirdly, the sample size was small due to the circumstances in which it was decided to evaluate this group and because the mode of delivery changed part-way through. This limits the confidence in quantitative impacts, and means it is unclear whether singers experienced benefits during the face-to-face or online part of the programme, or a combination of the two. Nevertheless, as a convenience pilot study, it provides useful indications of impacts, as well as informing future research. Similarly, the sub-optimal attendance during online sessions limits the extent to which impact can be assessed, although when combined with the interview feedback this provides useful information regarding barriers and facilitators to participation that can be addressed in both practice and future research. Finally, it is important to consider the context in which this trial took place. The developing COVID-19 pandemic was a considerable source of concern for many people with COPD, who were identified as being at an increased risk of severe COVID-19 or death. Though all the participants lived in London, the situation and their response to it, is likely to have differed between individuals, which intern, may have shaped their experience of the intervention.

Despite the necessary, yet unusual, adaptations to the methods, this study has provided interesting and potentially useful results to inform the development of further research regarding online singing group delivery and research. These findings are useful for existing SLH groups who are moving to online delivery of previously in-person sessions. They also provide the some of the first research findings to support the delivery of participatory online arts-in-health interventions in the context of COVID-19 related physical distancing.

The findings may also provide relevant insight for other related activities making an online transition such as pulmonary rehabilitation and Tai-Chi^34^, and dance for people with long-term medical conditions. Many of these activities had begun to develop and test online delivery approaches prior to the COVID-19 pandemic^35 36^, though the importance and potential utility of online delivery has now clearly increased^16^.

Further research should include larger studies assessing the health and wellbeing impact of online group singing in patient groups and for the wider population. Larger studies of SLH specifically, both online, and face-to-face (when appropriate to do so) remain a priority. Even after the acute phase of the COVID-19 pandemic online delivery of singing groups present an opportunity to widen access to certain groups of people. In-depth qualitative research exploring participant experience would also be valuable.

## Conclusions

In conclusion, this study suggests that group singing sessions that have had to change delivery from face-to-face to online may produce clinically significant impacts on depression scores, anxiety, and improve balance confidence in people with COPD. The findings also identify important differences between online and face-to-face delivery including technological barriers for online, and overcoming physical access barriers of face-to-face. Despite a general preference for face-to-face sessions, online delivery was still felt to provide substantial health and wellbeing benefits. Future research on digitally delivered singing groups is required.

## Data Availability

Data are available upon reasonable request.

## Acknowledgements

We would like to thank the Royal Brompton and Harefield Hospital Arts Team (RB&H Arts), and the study participants.

## Author Contributions

The authors meet criteria for authorship as recommended by the International Committee of Medical Journal Editors. All the authors played a role in the content and writing of the manuscript. In addition, KP performed the analysis of the data, NSH and KP prepared the first draft.

## Funding and Support

KP was supported by National Institute for Health Research Academic Clinical Fellowship award and the Imperial College Clinician Investigator Scholarship. The funders had no say in the design and conduct of the study; collection, management, analysis, and interpretation of the data; preparation, review, or approval of the manuscript; and decision to submit the manuscript for publication. This publication presents independent research. The views expressed are those of the authors and not necessarily those of the NHS, the NIHR or the Department of Health and Social Care.

## Licence for Publication

The Corresponding Author has the right to grant on behalf of all authors and does grant on behalf of all authors, an exclusive licence (or non exclusive for government employees) on a worldwide basis to the BMJ Publishing Group Ltd to permit this article (if accepted) to be published in BMJ Open Respiratory Research and any other BMJPGL products and sublicences such use and exploit all subsidiary rights, as set out in our licence (http://group.bmj.com/products/journals/instructions-for-authors/licence-forms).

## Data Availability Statement

Data are available upon reasonable request.

## Competing Interest

None declared.

## Competing interests

The authors have no conflicts of interest relevant to the content of this article.

## Notes

### Competing Interest Statement

The authors have declared no competing interest.

### Clinical Trial

ClinicalTrials.gov Identifier: NCT04034212

### Author Declarations

Ethical approval for this studywas granted by the National Health Service Health Research Authority, Stanmore REC (19/LO/0418).

